# The Spatial Immune Landscape of Mismatch Repair Deficient Endometrial Cancer: Implications for Clinical Outcomes

**DOI:** 10.64898/2026.03.12.26348249

**Authors:** Michelle Hollenberg, Catherine Hermann, Aliza Siegman, Wenke Liu, Cynthia Loomis, Valeria Mezzano, Shanmugapriya Selvaraj, Jimin Tan, Tianxiao Zhao, Joshua M. Wang, Lizabeth Katsnelson, Linda Procell, Esther Adler, Leslie Boyd, David Fenyö

**Author notes:** Correspondences (E.A.), (L.B.), (D.F.). Declaration of Competing Interests: The authors declare that they have no conflicts of interest.

## Abstract

Mismatch repair–deficient (dMMR) endometrial tumors are often responsive to immune checkpoint inhibitors (ICI), yet recurrence and variable treatment responses remain significant clinical challenges. Characterization of the tumor microenvironment, including immune cell composition and spatial organization, may reveal predictors of recurrence and ICI responsiveness. We performed multiplex immunofluorescence imaging on 16 dMMR endometrial tumors using a 36-antibody panel. Cells were segmented, phenotyped by unsupervised clustering, and analyzed to quantify cell type proportions and spatial relationships among intratumoral, peritumoral, and whole-tissue cell populations across clinical groups. Non-recurrent tumors (n = 10) exhibited higher intratumoral CD8⁺ T-cell proportions, tumor cell enrichment around CD8^+^ T cells, CD8^+^/CD4^+^ ratios, and PD-1^Low^ CD4⁺ T-cell proportions. In contrast, recurrent tumors (n = 6) showed higher CD4^+^ T cell proportions and endothelial cell enrichment surrounding CD8⁺ and PD-1⁺ CD8⁺ T cells. Among the recurrent tumors, compared to non-responders (n = 2), ICI responders (n = 4) had a higher proportion of PD-1^+^Ki67^+^ CD8^+^ T cells. Macrophage spatial organization also differed; non-responders displayed separate clusters of CD163⁺ macrophages and CD163⁻ macrophages, whereas responders demonstrated more dispersed macrophages co-localized with PD-1^+^ CD8⁺ T cells. Overall, these findings suggest that both immune cell composition and spatial arrangement are factors that contribute to recurrence and ICI response in dMMR endometrial cancer. Spatial profiling of the tumor microenvironment may provide biomarkers to guide patient stratification and precision immunotherapy strategies.

## INTRODUCTION

Endometrial cancer is the most common gynecologic malignancy in the United States, with rising incidence and mortality rates (1). The Cancer Genome Atlas identified four distinct molecular subtypes of endometrial cancer: Polymerase Epsilon ultra-mutated; microsatellite instability-high (MSI-H); copy-number high; and copy-number low (2). These molecular subtypes provide improved prognostic stratification compared with prior histology-based systems, and increasingly inform treatment selection (3–5).

Among 30 human cancer types, endometrial cancer has the highest prevalence of mismatch repair deficiency (dMMR), present in approximately 30% of cases (6). This molecular subtype is highly immunogenic, with high tumor mutational burden and neoantigen generation, promoting tumor-infiltrating lymphocytes (7–9), and is a strong predictor of response to immune checkpoint inhibition (ICI), as demonstrated in recent trials of advanced and recurrent disease (10–12). Despite this, nearly half of dMMR tumors fail to respond to ICI therapy, and dMMR tumors represent a substantial fraction of recurrent cases (13–30%), underscoring the need to understand the biological mechanisms driving both recurrence and therapeutic response in this endometrial cancer subtype (13). Persistent recurrence and variable response to treatment with ICI, despite heightened immune infiltration, suggest that additional factors of the TME contribute to disease progression.

The tumor microenvironment (TME), including the cellular composition, functional cell states, and spatial organization, plays a critical role in tumor biology, progression, and therapeutic response (1,2). Significant advances in spatial profiling technologies, such as multiplexed immunofluorescence (mIF) imaging, have enabled high-dimensional characterization of immune architecture at single-cell resolution (3,4). Although dMMR endometrial tumors are considered an immunologically and clinically favorable subtype, the heterogeneity within this population remains incompletely characterized (14).

In this pilot study, we evaluated the feasibility of multiplexed spatial imaging using the Akoya Biosciences PhenoCycler platform to characterize the immune microenvironment of dMMR endometrial tumors, and to identify cellular and spatial features associated with recurrence and response to immune checkpoint therapy.

## MATERIALS AND METHODS

### Study approval

This study was approved by the New York University Institutional Review Board.

### Patient cohort and tumor acquisition

The study was approved for up to 20 patients. The panel was planned to be divided in half between recurrent and non-recurrent tumors. The recurrent tumors were then to be divided in half between those that responded to ICI and those that did not. Patients with MSI-H tumors were retrospectively identified from an IRB and HIPAA-compliant database run by the NYU Langone Health Gynecologic Pathology team. Inclusion criteria encompassed patients with primary non-Lynch MSI-H tumors diagnosed between 2017 and 2021 with at least two years of follow up from diagnosis and who had information pertaining to treatment available in the electronic medical record (EMR). Clinical data was collected from the EMR, including demographic information, cancer data including staging and pathology, adjuvant treatment, recurrence, and treatment for recurrence (**Table 1**). Tumors were staged using the 2009 FIGO staging schema. Tumor tissue for the identified patients was obtained from the NYU Center for Biospecimen Research and Development, which stores all surgical tissue blocks obtained at NYU Langone Health. Tumor blocks were sectioned to 4 microns. Additional adjacent sections for hematoxylin and eosin (H&E) staining were taken.

**Table 1:** Patient Clinical Data.

|  | Non-recurrent<br>(n = 10) | Recurrent<br>(n = 6) | Recurrent<br>(n = 6) |  |
| --- | --- | --- | --- | --- |
|  |  |  | ICI non-responder<br>(n = 2) | ICI responder<br>(n = 4) |
| Age, years (range) | 73 (50-78) | 68.7 (48-77) | 73 (72-74) | 66.5 (48 - 77) |
| Stage (%) |  |  |  |  |
| 1A | 6 (60) | 1 (16.6) | 1 (50) | 0 (0) |
| 1B | 2 (20) | 1 (16.6) | 0 (0) | 1 (25) |
| IIIA | 1 (10) | 0 (0) | 0 (0) | 0 (0) |
| IIIC1 | 1 (10) | 3 (50) | 1 (50) | 2 (50) |
| IVB | 0 (0) | 1 (16.6) | 0 (0) | 1 (25) |
| Grade (%) |  |  |  |  |
| 1 | 7 (70) | 4 (66.7) | 1 (50) | 3 (75) |
| 2 | 3 (30) | 2 (33.3) | 1 (50) | 1 (25) |
| 3 | 0 (0) | 0 (0) | 0 (0) | 0 (0) |
| Adjuvant Therapy (%) |  |  |  |  |
| Yes | 4 (40) | 4 (66.7) | 1 (50) | 3 (75) |
| No | 6 (60) | 2 (33.3) | 1 (50) | 1 (25) |
| Adjuvant Radiation Therapy (%) |  |  |  |  |
| Yes | 3 (30) | 2 (33.3) | 1 (50) | 1 (25) |
| No | 7 (70) | 4 (66.7) | 1 (50) | 3 (75) |
| Adjuvant Chemo (%) |  |  |  |  |
| Yes | 2 (20) | 4 (66.7) | 1 (50) | 3 (75) |
| No | 8 (80) | 2 (33.3) | 1 (50) | 1 (25) |
| BMI, kg/m<sup>2</sup> (range) | 28.3 (21-37) | 32.3 (18-46) | 34 (28-40) | 31.5 (18-46) |
| Race (%) |  |  |  |  |
| White | 6 (60) | 5 (83.3) | 2 (100) | 3 (75) |
| Black | 3 (30) | 0 (0) | 0 (0) | 0 (0) |
| Other | 1 (10) | 1 (16.6) | 0 (0) | 1 (25) |
| dMMR gene loss (%) |  |  |  |  |
| MLH1, PMS2 | 6 (60) | 6 (100) | 2 (100) | 4 (100) |
| MSH6 | 2 (20) | 0 (0) | 0 (0) | 0 (0) |
| MSH2, MSH6 | 1 (10) | 0 (0) | 0 (0) | 0 (0) |
| PMS2 | 1 (10) | 0 (0) | 0 (0) | 0 (0) |
| MLH1 promoter<br>hypermethylation (%) |  |  |  |  |
| Yes | 6 (60) | 6 (100) | 2 (100) | 4 (100) |
| NA | 4 (40) | 0 (0) | 0 (0) | 0 (0) |
| Smoking Status (%) |  |  |  |  |
| Yes | 3 (30) | 4 (66.7) | 0 (0) | 4 (100) |
| No | 7 (70) | 2 (33.3) | 2 (100) | 0 (0) |

Clinical features of 16 dMMR endometrial cancer. Data presented as mean (range) or count (percentage). ICI = immune checkpoint inhibition, Age = age at time of surgery, BMI = Body Mass Index, dMMR = Mismatch Repair Deficiency.

### Antibody panel design

Biomarkers chosen for PhenoCycler analysis were selected based on having a known role in endometrial cancer pathogenesis and the proteomic analysis of endometrial cancer (15,16). In addition, immune, standard nuclear, and epithelial markers were included. For the full antibody panel, including clones, manufacturers, and catalog numbers, please see **Supplementary Table 1**. Previously validated and conjugated antibodies that were available from Akoya Biosciences were purchased directly from Akoya Biosciences®. Antibodies that were not available were ordered from outside sources and validated by the NYU Langone Health Experimental Pathology lab. Once the antibodies were validated, they were custom conjugated by Akoya Bioscience to oligonucleotide barcodes to allow for their use with PhenoCycler.

### Multiplexed immunofluorescence

Co-detection by indexing (CODEX) was performed on a PhenoCycler Fusion® instrument (Akoya Biosciences), according to the manufacturer’s instructions. In brief, 5 µm thick tissue sections were placed on superfrost plus slides and baked overnight at 60 C prior to deparaffinization through a series of xylenes and ethanol washes. Antigen retrieval was performed in an Instant Pot® pressure cooker with 1x AR9 (Cat# AR900250ML, Akoya Biosciences) for 20 min. at high pressure. Slides were cooled, placed in Akoya hydration buffer (Cat#240196, Akoya Biosciences) for 2 min, followed by Akoya staining buffer (Cat#240198, Akoya Biosciences) for 20 min. Slides were then incubated with the panel of oligo-linked antibodies for 3 hours at room temperature. After washing, slides were fixed 1.6% PFA (16% PFA from Electron Microscopy Sciences, Cat# 15710) in Akoya storage buffer (Cat#240197, Akoya Biosciences) for 10 mins at room temperature followed by chilled 100% methanol at 4 C for 5 minutes and then Akoya fixative reagent (Cat#23112, Akoya Biosciences) diluted in PBS for 20 min at RT. Slides were kept in Akoya storage buffer (Cat#240197, Akoya Biosciences) at 4 C until ready to image. Reporter plates containing complementary oligonucleotide conjugated fluorophores were prepared according to panel design and slides applied to the microfluidics flow chamber (**Supplemental Table 1**). Automated protein detection and imaging were performed at 0.51 px/mm (20X) on the PhenoCycler-Fusion System (Akoya Biosciences). Oligo-linked reporter dyes from each well were iteratively applied to the slide and imaged, followed by removal of the oligo-linked fluorophores before moving on to the next cycle. The images from each cycle were then compiled into a single QPTIFF file with Akoya Fusion software (1.0.8), and the scans reviewed with Phenochart Viewer 2.2.0 to ensure appropriate staining before uploading to the NYU Grossman School of Medicine OMERO Plus image data management system (Glencoe Software).

### Image preprocessing

Multiplex Immunofluorescence (mIF) and matched H&E images were processed in their original formats (.qptiff, .svs). Each pixel corresponded to 0.5 µm. Tumor regions were annotated by a pathologist on the H&E slides and mapped to the corresponding mIF whole-slide images using the Valis registration pipeline (17). Within tumor regions, tissue artifacts (e.g., folds and foreign material) were manually annotated and excluded using OMERO Plus and Pathviewer(r) (Glencoe Software) (**Supplementary Fig. S1A**). Necrotic areas were annotated by a pathologist and removed from downstream analysis.

### Cellular segmentation

Individual cells were identified using the Mesmer model from DeepCell, which supports both nuclear and whole-cell segmentation (18). Whole-slide images were divided into tiles for cell segmentation (128 µm x 128 µm). Tiles were assigned to nuclear (DAPI-based) or whole-cell segmentation according to E-cadherin expression, which served as a consistent membrane marker. Segmentation type was determined per tile by calculating the mean intensity of the top 5% of E-cadherin–positive pixels. Tiles were then clustered within each sample using unsupervised k-means clustering (k = 3). The 2 clusters with the highest centroids underwent whole-cell segmentation, whereas the remaining cluster underwent nuclear segmentation with a 2-pixel (1 µm) expansion to approximate cell boundaries. An example tissue sample showing the assigned segmentation types across all tiles is shown, along with one representative raw and annotated tile from each segmentation type **(Supplementary Fig. S1A-B)**. Segmentation masks from each tile were subsequently mapped back to whole-slide coordinates for all downstream analyses. Mean marker intensities per cell were calculated.

### Cellular quality control and normalization prior to lineage marker cell phenotyping

Cells with low mean DAPI intensity, indicative of incomplete nuclei, artifacts or autofluorescence, were excluded from analysis. For each lineage marker, to distinguish true biological signal from background or autofluorescence, we applied k-means clustering (k = 3) to the per-cell mean intensities within each sample. Clusters corresponding to background or oversaturation were identified by visual inspection in Omero, and the mean cell values in those clusters were set to zero. When clustering did not clearly separate background or oversaturated artifacts, a manual lower-intensity or upper-intensity cutoff was applied. Representative examples of background thresholding for CD4 and CD68 are shown (**Fig. S1C**).

### Lineage marker cell phenotyping

Within each sample, lineage marker intensities were capped at the 99.9^th^ percentile then normalized using an arcsinh transformation and z-score scaling. This approach best stabilizes variance, compresses extreme values, and standardizes marker distributions prior to unsupervised clustering for lineage cell phenotyping (19,20). Normalized lineage marker intensities were then clustered using unsupervised k-means clustering (k = 27) to capture all expected immune and stromal populations. The optimal number of clusters was determined using the elbow method, selecting the point where additional clusters no longer substantially reduce within-cluster variance. Each cluster was annotated based on its dominant marker, informed by canonical expression patterns and visual inspection in Omero (**Fig. S1D**). Heterogeneous clusters, which either lacked a single dominant marker or contained multiple high-expressing markers, were further refined. Marker intensity thresholds were determined per sample using OMERO to distinguish true biological signal from background. Cells exceeding a single lineage marker threshold were assigned to that marker’s cell type; those exceeding multiple thresholds were retained as heterogeneous.

Residual heterogeneous cells were then assigned using a percentile-based comparison to reference populations. For each lineage cell type, the distribution of marker intensities among previously assigned non-heterogeneous cells was calculated to establish the reference distribution of marker expression. Marker intensities of heterogeneous cells were compared to these distributions to determine whether their expression was consistent with a given cell type. Cells whose marker intensity fell within the reference distribution of a single lineage were assigned to that lineage. Cells whose marker intensities were consistent with more than one lineage were labeled as a mixed cell type population and cells that did not fall within the expected range of any lineage were classified as undefined stromal cells.

### Functional marker cell phenotyping

For non-lineage markers, per-cell marker intensities were analyzed on a per-marker, per-lineage cell type basis (e.g., PD-1 expression within CD8⁺ T cells). Within each sample and cell type, k-means clustering (k = 4-6) was applied to separate background from true signal. Clusters corresponding to background staining were identified by visual inspection in OMERO and set to zero. To further remove artifacts, clusters containing fewer than 15 cells were considered spurious and excluded from analysis. The remaining cells were further classified into low and high expression groups using Gaussian mixture modeling (2 components) (5). This approach generated low and high expression categories for each marker per cell type, while positive expression included all cells identified as either low or high.

### Intratumoral, peritumoral, and whole tissue cell classification

We performed downstream analyses by regional assignment (intratumoral versus peritumoral) and across the whole tissue, capturing distinct immune compositions and improving prediction of clinical outcomes, including recurrence risk (21,22). Cells were classified as intratumoral or peritumoral based on E-cadherin expression, which served as a proxy for tumor cell localization and was confirmed by visual inspection of the images. Assignments were made using k-means clustering (k = 3) on per-cell mean E-cadherin intensities. Clusters designated as intratumoral corresponded to cells within or immediately adjacent to tumor nests, whereas peritumoral clusters corresponded to cells in the surrounding stroma between tumor cells. For samples in which k-means clustering did not fully capture tumor boundaries, E-cadherin thresholds were manually determined and validated using OMERO Plus and PathViewer to distinguish intratumoral from peritumoral cells. An example of these cell level regional assignments is shown in **Fig. 1A**. All proportion and spatial interaction analyses were then performed based on these regional assignments, as well as across the whole tissue (**Fig. 1B)**.

**Figure 1.**
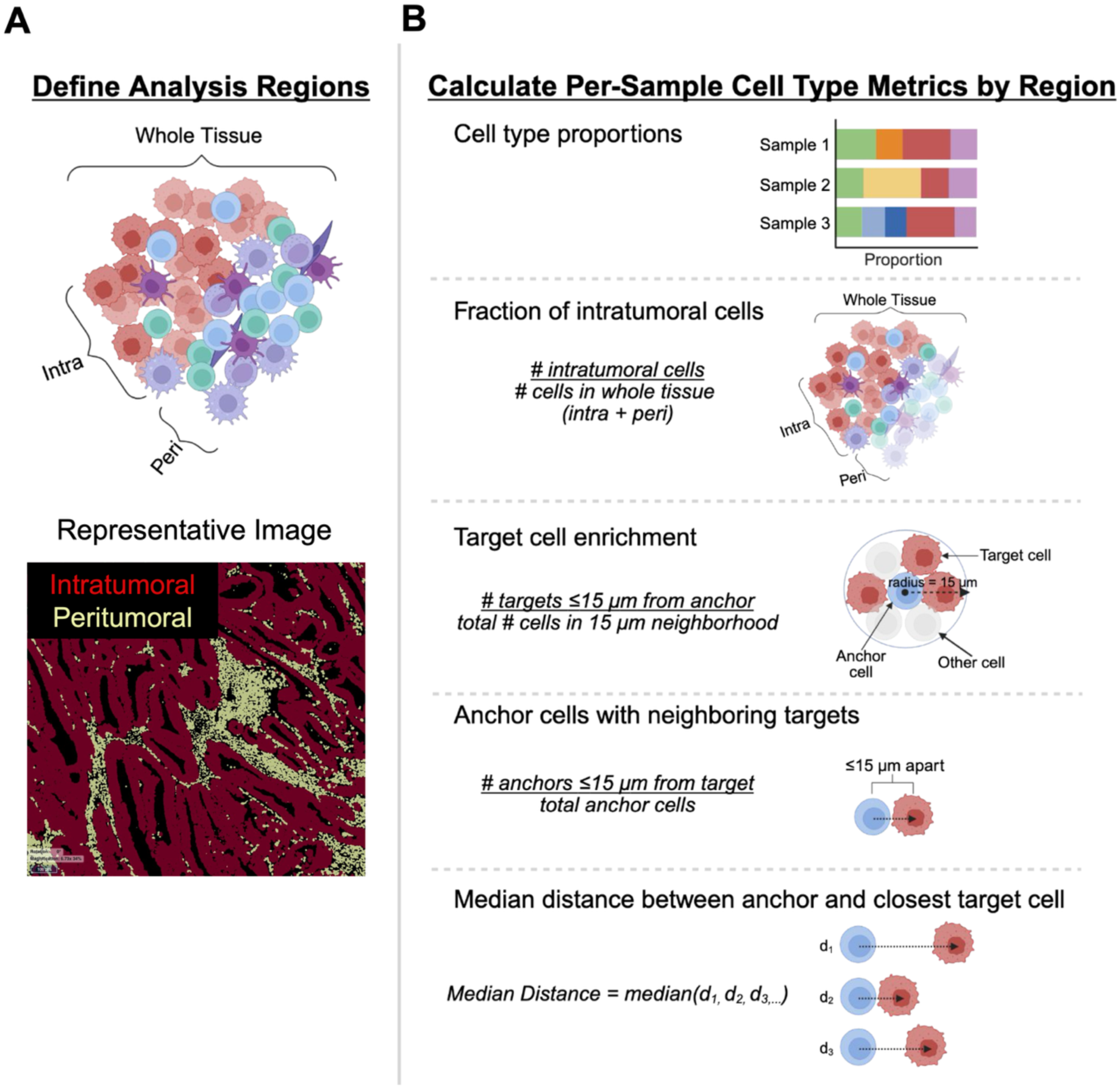
Regional cell type assignment, proportion, and spatial analysis. (A) Cells were classified as intratumoral or peritumoral based on relative E-cadherin expression. Whole tissue analyses included both regions. Top: schematic. Bottom: representative image showing cells annotated by regional assignment. (B) For each region and sample, multiple metrics were calculated, including cell type proportions, fraction of intratumoral cells, and spatial relationships between anchor and target cells. Per sample metrics were grouped by clinical outcome and summarized with boxplots. Statistical comparisons used Mann–Whitney tests for recurrence groups (*p* < 0.05) and effect size for response groups.

### Cell type proportion analysis

Proportions were calculated relative to the appropriate parent population, which varied by cell type and marker (**Fig. 1B**). For non-T cell populations, the proportion was calculated as the number of cells of interest divided by the total number of cells (including defined, mixed, and undefined cells). For T cell populations, proportions were calculated relative to the total T-cell population (CD8^+^, CD4^+^, and CD4^-^ CD8^-^ CD3^+^ T cells), rather than to all cells, to assess differences in T-cell subset composition. Marker-defined T-cell subpopulations, such as Ki67⁺ CD8⁺ T cells, were quantified as the number of marker-positive cells divided by the total cells in the parent population which includes Ki67^+^ and Ki67^-^ CD8^+^ T cells. For analyses of high- or low-expressing cell types, such as PD1^Low^ CD4^+^ T cells, proportions were calculated either relative to the total number of marker-positive cells (PD1^+^ CD4^+^ T cells) or relative to the total number of parent cells (CD4^+^ T cells), depending on the biological question being addressed. For endothelial cells, rather than relying on cell counts, vasculature was quantified using HALO to measure CD31^+^ area, providing a biologically meaningful measure of vasculature coverage that accounts for their irregular morphology. The CD31⁺ area (mm²) was divided by the total tissue area (mm²) to calculate the fraction of tissue occupied by vasculature.

### Scimap interaction analysis

We used scimap, a computational package for spatial analysis of multiplexed images, to quantify the local composition of target cells around anchor cells (23). Specifically, we applied the *spatial_count()* function with the radius method, which identifies neighbors within a specified distance for each cell. To approximate the distance at which cells are likely to physically interact, we set the neighborhood radius to 15 µm. A neighborhood includes the central anchor cell, target cells, and all other cells within the defined radius. Analyses were performed using the output from spatial_count(), and results were compared across clinical groups.

To calculate the target cell enrichment, for each anchor cell with at least 1 target cell within its neighborhood, we computed the proportion of target cells relative to the total number of cells in this neighborhood. We then calculated the mean of these proportions per sample (**Fig. 1B**).

To calculate the proportion of anchor cells with neighboring targets, we calculated the proportion of anchor cells that had at least one neighboring cell of a specified target cell type within a 15 µm radius relative to total anchor cells in the sample (**Fig. 1B**).

### Median distance analysis

To complement the neighborhood-based analyses, we quantified spatial relationships between anchor and target cell types without a fixed interaction radius. For every anchor–target cell type pair, the distance from each anchor cell to its nearest target cell was computed using *cKDTree* from *scipy.spatial*, which allows efficient nearest-neighbor queries. The nearest neighbor (k = 1) was used when the anchor and target cell types were different, whereas the second-nearest neighbor (k = 2) was used when the anchor and target cell types were the same. This prevents the anchor cell from being counted as its own neighbor. For each anchor–target pair, the median nearest-neighbor distance was calculated to summarize the typical spatial separation between the cell types (**Fig. 1B**). These median distances were then compared across clinical groups to identify anchor–target cell type pairs with altered spatial proximity.

### Statistical Analysis

Statistical and computational analyses were performed in Python (version 3.11.8). Group differences were assessed using the Mann-Whitney U test for nonparametric data. Statistical significance was set at *P < 0.05*. For analyses involving small sample sizes, specifically the recurrent cohort stratified by ICI responders and non-responders, formal hypothesis testing was not performed. Instead, median differences were reported as an effect size to avoid unstable *P* values and to provide a descriptive measure of group differences. Interpretation focused on comparisons demonstrating minimal overlap between the responder and non-responder groups. Boxplots show the median (center line) and interquartile range (IQR; box), with whiskers extending to 1.5× the IQR.

### Image-based validation of marker assignments

All cell segmentation, phenotyping, and spatial analyses were visually validated using OMERO Plus (Glencoe Software). Segmented cell masks and associated metadata (e.g., cell types, marker intensities, cluster assignments) were overlaid on the original mIF images to inspect marker expression patterns, confirm cell phenotypes, and assess spatial neighborhoods. Paired H&E images were used to guide region-of-interest selection and verify tissue architecture. This interactive visualization ensured robust functional marker assignment, accurate cell segmentation, and minimized technical artifacts across the dataset.

## RESULTS

### Clinical cohort and immune landscape of dMMR endometrial tumors

Sixteen dMMR endometrial patients meeting inclusion criteria were identified and had tumor tissue available in the repository. Tumors were categorized by recurrence with at least 2 years of follow-up after first-line surgical treatment into non-recurrent (n = 10) and recurrent (n = 6) groups. Among recurrent cases who received ICI therapy, response to anti-PD-1 ICI therapy (pembrolizumab or dostarlimab) was assessed, with responders (n = 4) achieving disease control for ≥ 6 months after treatment initiation and non-responders (n = 2) showing progression within 6 months. Clinical groups were stage and grade matched to the extent possible (**Fig. 2A**, **Table 1**).

**Figure 2.**
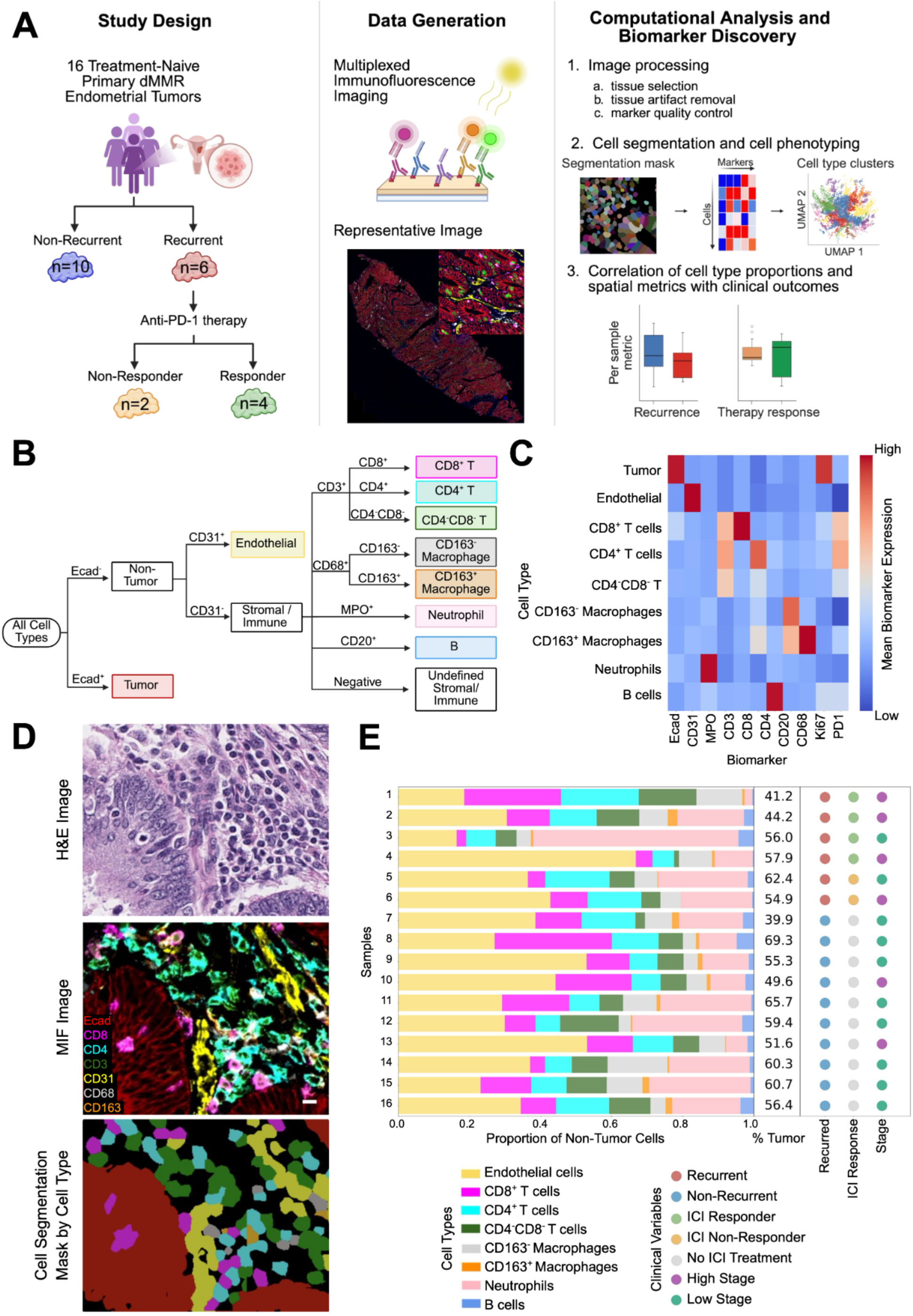
Experimental overview and validation of single-cell phenotyping in dMMR endometrial tumors. (A) Project overview including study design, data generation, and computational analysis. (B) Schematic of the hierarchical decision tree used to annotate cell types. (C) Heatmap showing mean marker expression per cell type after final assignments (see cluster level results in **Supplementary Fig. S1D**). Expression values are z-scored per marker to better visualize the dominant protein expressed per cell type. Blue indicates low and red indicates high normalized mean expression. (D) Validation of cell-type assignments by comparing H&E staining, multiplexed immunofluorescence (mIF), and the corresponding cell type masks. (E) Proportions of non-tumor cell types are shown relative to all non-tumor whole tissue cells; % Tumor reflects tumor cells as a fraction of total whole tissue cells across all samples. Clinical variables (recurrence, ICI response, and stage) are indicated.

Across all images, we identified 11,117,631 cells, of which 11,100,415 were well-defined, 17,216 exhibited mixed lineage marker expression, and 3,030,735 were undefined. Undefined cells lacked detectable expression of lineage markers, likely reflecting markers outside our panel or technical limitations, whereas mixed-phenotype cells expressed more than one lineage marker. Analysis of major populations revealed CD8⁺, CD4⁺, and CD3⁺ T cells, B cells, macrophages, endothelial cells, and tumor cells, with hierarchical classification illustrated in **Fig. 2B** and canonical marker expression validating each population shown in **Fig. 2C**.

Functional marker expression provided insight into cellular states. Ki67 expression, indicating proliferative activity, was highest in tumor cells, as well as T and B cells, whereas PD-1 expression, indicating recent antigen engagement, was enriched in T cells. Together, these markers enable assessment of both cellular composition and functional immune states, which were subsequently evaluated in relation to recurrence and response to immunotherapy. Cell type assignments were validated by comparing H&E morphology, mIF signals, and segmentation labels (**Fig. 2D**).

Proportions of non-tumor cells and % Tumor varied markedly across samples (**Fig. 2E**), reflecting substantial intertumoral heterogeneity in immune composition and motivating comparisons of cell-type distributions across clinical outcomes in subsequent analyses.

### Non-recurrent tumors exhibit higher intratumoral CD8⁺ T-cell proportions and closer CD8–tumor proximity

To evaluate region-specific CD8⁺ T-cell distributions, we quantified CD8⁺ T cells in intratumoral, peritumoral, and whole-tissue regions. Non-recurrent tumors demonstrated a higher proportion of intratumoral CD8⁺ T cells among total T cells compared to recurrent tumors (p = 0.0225; **Fig. 3A–B**, top), with a similar trend that did not reach significance when calculated relative to total cells (**Supplementary Fig. S2A**) or among peritumoral or whole tissue cells (**Supplementary Fig. S2B**). Interestingly, the only two low-stage tumors that recurred had particularly low intratumoral CD8⁺ T-cell proportions, suggesting that reduced CD8⁺ T cell infiltration may outweigh the otherwise favorable prognosis associated with low stage (**Supplementary Fig. S2C**).

**Figure 3.**
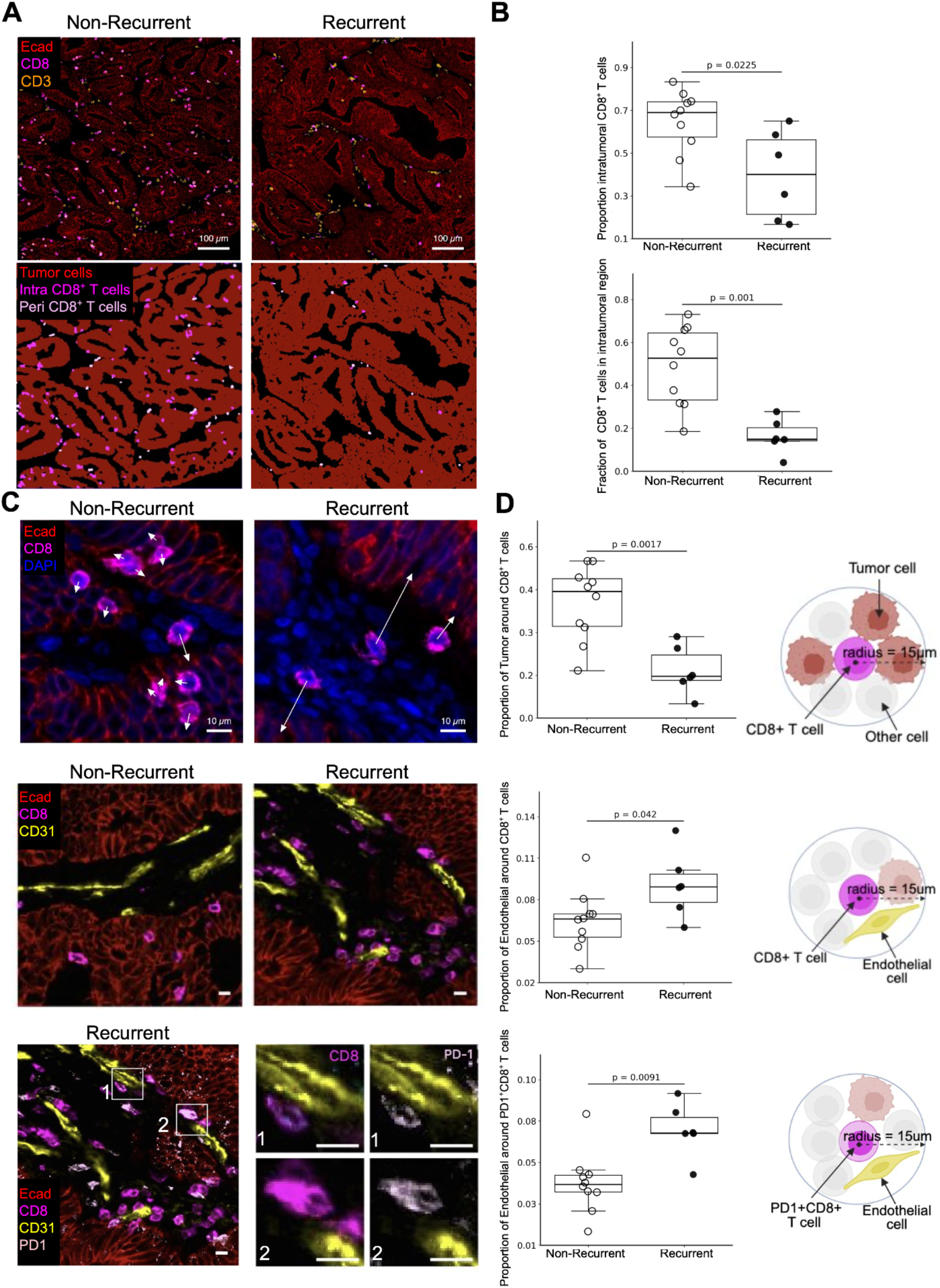
CD8^+^ T cell distribution and surrounding neighborhood enrichment differ between non-recurrent (N=10) and recurrent (N=6) tumors. (A) Top: representative multiplexed immunofluorescence images showing CD8⁺ T cells (CD8, fuchsia) and all T cells (CD3, orange); tumor epithelial cells are marked by E-cadherin (Ecad, red). Bottom: cell masks differentiate intratumoral (light pink) and peritumoral (dark pink) CD8^+^ T cells, with more intratumoral than peritumoral CD8^+^ T cells in the non-recurrent tumor. Scale bars = 100 µm. (B) Top: proportion of intratumoral CD8⁺ T cells of total T cells higher in non-recurrent tumors, calculated as the number of intratumoral CD8⁺ T cells divided by the total number of intratumoral T cells per sample. Bottom: proportion of CD8⁺ T cells located intratumorally across the whole tissue calculated as the number of intratumoral CD8⁺ T cells divided by the number of whole tissue (intratumoral + peritumoral) CD8⁺ T cells per sample. (C) Representative multiplexed immunofluorescence images showing neighborhood enrichment (≤15 µm). Top: tumor cells around CD8^+^ T cells; arrows highlight closer proximity in a non-recurrent tumor. Middle: endothelial cells (CD31, yellow) around CD8⁺ T cells, illustrating more endothelial enriched neighborhoods in a recurrent tumor. Bottom: endothelial cells around PD-1⁺ CD8⁺ T cells (PD-1, light pink) in a recurrent tumor. Boxes 1 and 2 highlight representative examples of endothelial cells within 15 µm of a PD-1⁺ CD8⁺ T cell. Scale bars = 10 µm. (D) For each sample, the mean proportion of tumor cells around CD8⁺ T cells (top), endothelial cells around CD8⁺ T cells (middle), and endothelial cells around a subset of CD8⁺ T cells that are PD-1⁺ (bottom); each within a 15 µm radius. Calculated as the number of surrounding tumor or endothelial cells divided by the total number of cells in the neighborhood. Boxplots show the mean proportion per sample. Schematics next to each plot illustrate the neighborhood definition. *P* < 0.05 by Mann–Whitney U test.

We next assessed the spatial distribution of CD8⁺ T cells between intratumoral and peritumoral regions by calculating the proportion of total CD8⁺ T cells localized to the intratumoral compartment. This proportion was significantly higher in non-recurrent tumors (p = 0.001; **Fig. 3A–B**, bottom), consistent with preferential intratumoral localization.

We next assessed CD8⁺–tumor spatial relationships across the whole tissue. Representative images showed shorter CD8⁺–tumor distances in non-recurrent tumors compared to recurrent tumors (**Fig. 3C**). To quantify this observation, we measured local tumor cell enrichment within a 15 µm radius of each CD8⁺ T cell, a distance chosen to approximate potential cell–cell interactions. Non-recurrent tumors exhibited a higher mean proportion of neighboring tumor cells within this radius (p = 0.0017; **Fig. 3D**). Similarly, a greater proportion of CD8⁺ T cells had at least one tumor cell within 15 µm, and the median nearest CD8–tumor distances were shorter in non-recurrent tumors (p = 0.016 & 0.0426, respectively; **Supplementary Fig. S2D**).

Together, these analyses demonstrate that non-recurrent tumors are characterized by both increased intratumoral CD8⁺ T cell enrichment and reduced CD8–tumor spatial separation, reflecting tighter spatial coupling between CD8⁺ T cells and tumor cells.

### Endothelial cells are enriched around CD8⁺ and PD1^+^ CD8^+^ T cells in recurrent tumors

To investigate potential mechanisms limiting CD8⁺ T-cell infiltration into the intratumoral region of recurrent tumors, we evaluated the role of tumor vasculature. The overall amount vasculature, measured as CD31⁺ tissue area relative to total tissue area, and the proportion of tumor cells within 15 µm of endothelial cells were similar between clinical groups (**Supplementary Fig. S2E**, left and right). This indicates that neither the abundance of vasculature nor its proximity to tumor cells accounts for the reduced CD8⁺ T-cell infiltration in recurrent tumors. Therefore, the spatial relationship between CD8⁺ T cells and individual endothelial cells was examined. Among all cell types analyzed, only endothelial cells showed a significant increase in neighborhood enrichment around CD8⁺ T cells in recurrent tumors, with a higher proportion located within 15 µm of each CD8⁺ T cell (p = 0.042; **Fig. 3C-D**, middle) and a shorter median nearest CD8–endothelial distance (p = 0.011; **Supplementary Fig. S2F**). These contacts were concentrated at the tumor edge and surrounding stroma, indicating that despite no differences in the amount of vasculature or tumor–endothelial proximity, CD8⁺ T cells remain close to endothelial cells rather than infiltrating the tumor core in recurrent tumors.

We next evaluated which functional CD8⁺ T-cell subsets contribute to these vascular associations in recurrent tumors. PD-1⁺ CD8⁺ T cells, representing antigen-experienced effector T cells, constituted a similar proportion of CD8⁺ T cells in recurrent and non-recurrent tumors (**Supplementary Fig. S2G**), but were preferentially located near endothelial cells in recurrent tumors. Specifically, PD-1⁺ CD8⁺ T cells exhibited a higher mean proportion of neighboring endothelial cells within 15 µm (p = 0.0091; **Fig. 3C-D**, bottom) and a shorter median nearest distance to endothelial cells (p = 0.0047; **Supplementary Fig. S2H**).

Together, these results indicate that while overall vasculature and tumor–endothelial proximity are similar between recurrent and non-recurrent tumors, CD8⁺ T cells, particularly the antigen-experienced PD-1⁺ subset, in recurrent tumors are preferentially retained near endothelial cells, whereas more CD8⁺ T cells infiltrate the intratumoral region in non-recurrent tumors. This spatial retention of a functionally relevant subset may limit engagement with tumor cells and help explain differences in antitumor immune activity.

### Although CD4⁺ T cells have a higher proportion of T cells in recurrent samples, the PD-1 low subset is more prevalent in non-recurrent tumors

Having characterized CD8⁺ T cell proportions, localization, and neighborhood enrichment, CD4⁺ T cells were analyzed to understand how other T cell populations associate with recurrence. CD4⁺ T cell enrichment was assessed relative to both CD8^+^ T cells and the total T cell population, with representative images shown **(Fig. 4A**). Non-recurrent tumors exhibited a higher intratumoral CD8⁺/CD4⁺ T cell ratio, reflecting CD8⁺ T cell dominance among this intratumoral population (p = 0.0312, **Fig. 4B**). In contrast, recurrent tumors showed a higher proportion of CD4⁺ T cells among all whole tissue and intratumoral T cells (p = 0.0196 and 0.042, respectively; **Fig. 4C**), with similar trends but reaching no significance in the peritumoral region (**Supplementary Fig. S3A**).

**Figure 4.**
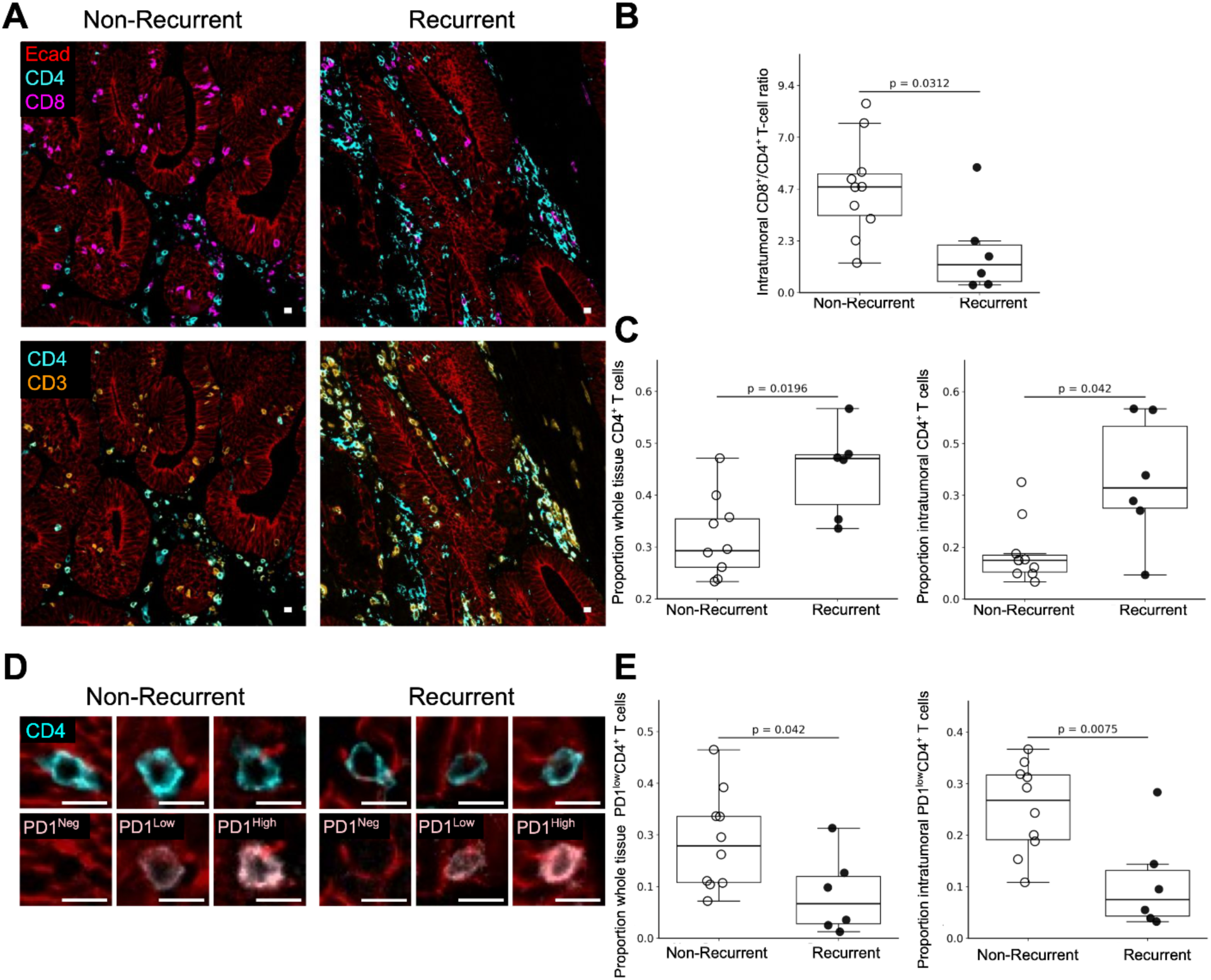
Recurrent tumors (n = 6) show CD4⁺ T cell enrichment, while PD-1^Low^CD4⁺ T cell subset are enriched in non-recurrent tumors (n = 10). (A) Representative multiplexed immunofluorescence images. Top: CD4⁺ T cells (teal) and CD8⁺ T cells (CD8, fuchsia). Bottom: CD4⁺ T cells and total T cells (CD3, orange). Tumor epithelial cells are marked by E-cadherin (Ecad, red). (B) Ratio of intratumoral CD8⁺ T cells to CD4⁺ T cells higher in non-recurrent tumors. (C) Whole tissue (left) and intratumoral (right) proportion of CD4⁺ T cells of total T cells are higher in recurrent tumors calculated as the number of intratumoral CD4⁺ T cells divided by the total number of intratumoral or whole tissue T cells per sample. (D) Representative multiplexed immunofluorescence images showing individual CD4⁺ T cells with negative (PD-1^Neg^), low (PD-1^Low^), and high (PD-1^High^) PD-1 expression, illustrating consistent expression assignments in non-recurrent and recurrent tumors (PD-1, light pink). (E) Higher whole tissue (left) and intratumoral (right) proportion of a subset of CD4⁺ T cells with low PD-1 expression in non-recurrent samples. The proportion of PD-1^Low^ CD4⁺ T cells was calculated as the number of CD4⁺ T cells with low PD-1 expression divided by the total number of CD4⁺ T cells, within each region. *P* < 0.05 by Mann–Whitney U test. Scale bars = 10 µm.

The functional state of CD4⁺ T cells via PD-1 expression was evaluated. Representative images confirmed consistent PD-1 labeling across PD-1^Negative^, PD-1^Low^, and PD-1^High^ subsets (**Fig. 4D**). Although recurrent tumors had a higher proportion of CD4⁺ T cells, non-recurrent tumors exhibited a higher proportion of whole tissue and intratumoral CD4⁺ T cells with low PD-1 expression (p = 0.042 and p = 0.0075, respectively; **Fig. 4E**), consistent with an activated rather than exhausted phenotype. The proportion of CD8^+^ PD-1^High^ T cells of total CD8^+^ T cells, indicative of a more exhausted phenotype, was similar across clinical groups (**Supplementary Fig. S3C**). In contrast to CD8⁺ T cells, CD4⁺ T cells maintained similar spatial relationships with tumor and endothelial cells across recurrence groups (**Supplementary Fig. S3D**).

Together, these analyses define the spatial distribution and functional states of T-cell subsets in recurrent and non-recurrent dMMR tumors. Non-recurrent tumors are defined by intratumoral CD8⁺ T-cell enrichment, closer CD8–tumor proximity, and an increased proportion of PD-1^Low^ CD4^+^ T cells, whereas recurrent tumors show increased CD8–endothelial associations particularly among PD-1⁺ CD8⁺ subsets and an increased relative enrichment of CD4⁺ T cells.

Together, these results indicate that recurrence is associated with altered T-cell composition and spatial organization, marked by diminished CD8⁺ tumor engagement and preferential retention near vasculature.

### Antigen-experienced, proliferative CD8⁺ T cells are enriched in ICI responders

In contrast to the recurrence-associated alterations in T-cell composition and spatial organization, ICI response was characterized by distinct CD8⁺ T-cell phenotypes. Overall CD8⁺ T-cell proportions were similar between groups (**Supplementary Fig. S4A**), prompting analysis of PD-1 expression to capture functional differences, as PD-1⁺ CD8⁺ T cells are central to anti–PD-1 responses. Using Gaussian mixture modeling, CD8^+^ T cells were classified as PD-1^Low^ and PD-1^High^ subsets, with representative cells shown in **Fig. 5A**. Responders had higher intratumoral proportions of PD-1^+^ CD8^+^ T cells among CD8^+^ T cells and PD-1^High^ CD8^+^ T cells among PD-1^+^ CD8^+^ T cells (effect size = 0.225 and 0.0667, respectively; **Fig. 5B**, left, right).

**Figure 5.**
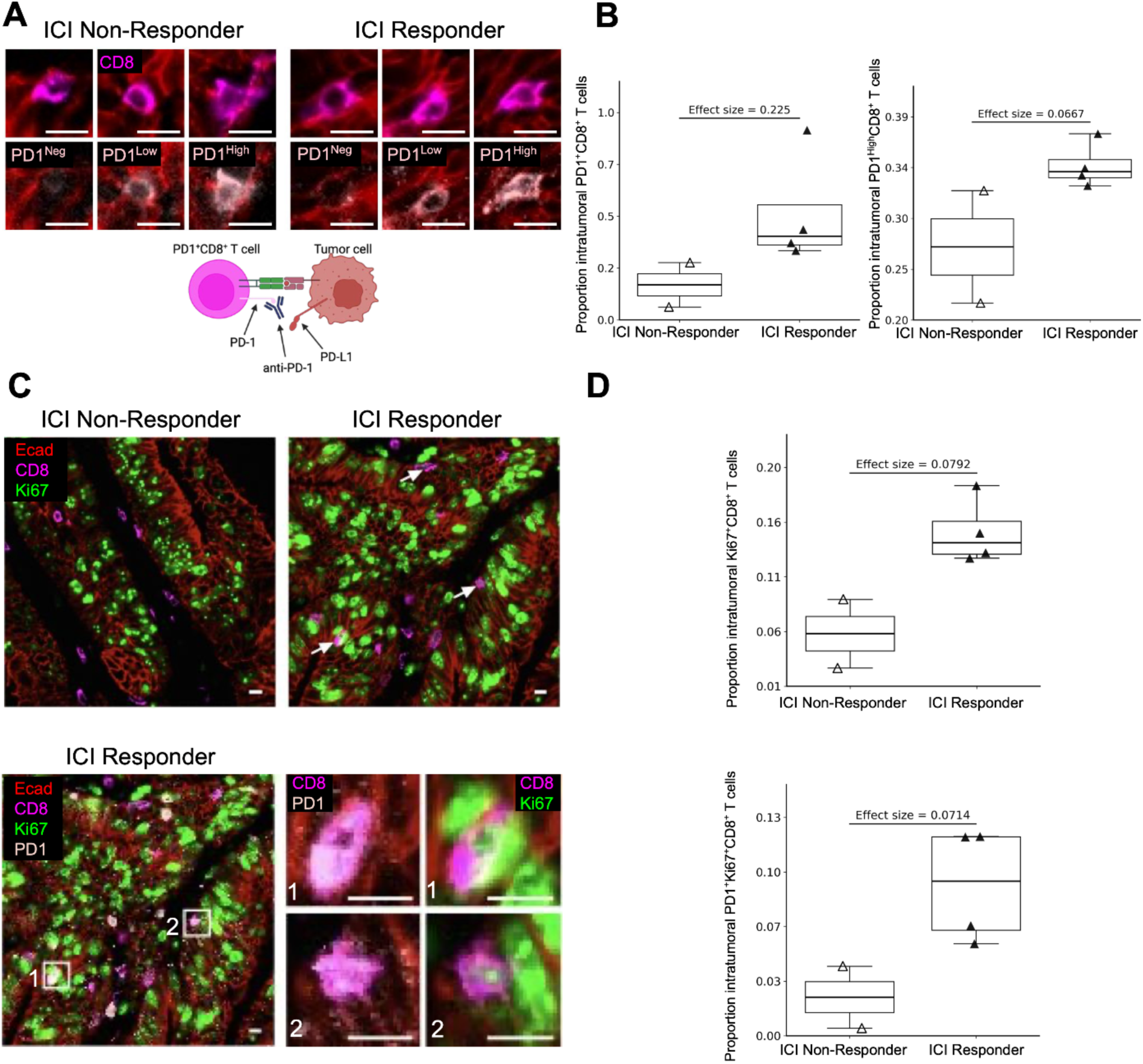
PD-1 and Ki67 expression in CD8⁺ T cells distinguish ICI responders. (A) Representative multiplexed immunofluorescence images showing individual CD8⁺ T cells (CD8, fuchsia) with negative (PD-1^Neg^), low (PD-1^Low^), and high (PD-1^High^) PD-1 (light pink; antigen-experience marker) expression in tumors from a non-responder and responder, illustrating consistent expression classification. Anti-PD-1 therapy schematic shown below. (B) Higher proportions of PD-1^+^ CD8^+^ T cells (left) and PD-1^High^ CD8^+^ T cells (right) observed in responders calculated by dividing PD-1^+^ CD8^+^ T cells by total CD8^+^ T cells and dividing PD-1^High^ CD8^+^ T cells by total PD-1^+^ CD8^+^ T cells. The PD-1^+^ CD8^+^ T cell population includes both PD-1^Low^ and PD-1^High^ cells. (C) Representative multiplexed immunofluorescence images of CD8⁺ T cells. Ki67⁺ cells (green; proliferation marker) are shown in an ICI non-responder and responder (top), while PD-1⁺Ki67⁺ cells are highlighted in an ICI responder (bottom). Top: white arrows indicate intratumoral Ki67^+^ CD8^+^ T cells in responder with no co-expressed cells shown in the non-responder image. Bottom: white boxes highlight intratumoral PD-1⁺Ki67⁺ CD8⁺ T cells, corresponding to single-cell regions of co-expression. (D) Proportion of intratumoral Ki67⁺ CD8⁺ T cells (top) and PD-1^+^Ki67⁺ CD8⁺ T cells (bottom) relative to the total number of intratumoral CD8⁺ T cells. *P* < 0.05 by Mann–Whitney U test. Scale bars = 10 µm.

We next assessed CD8⁺ T-cell proliferation using Ki67, a marker of actively cycling cells that occurs following activation, to complement the functional insights provided by PD-1 expression. Responding tumors had higher intratumoral Ki67⁺ CD8⁺ T cell proportions (effect size = 0.0792; **Fig. 5C–D**, top), consistent with enhanced intratumoral proliferation in effective anti-tumor responses.

Consistent with these findings, a higher proportion of CD8^+^ T cells with the co-expression of PD-1 and Ki67 was found in responders (effect size = 0.0714, **Fig. 5C-D**, bottom). Trends in other regions are shown in **Supplementary Fig. S4D-G.**

Together, these findings highlight distinct aspects of CD8⁺ T-cell biology associated with therapeutic response, linking antigen experience and proliferative capacity with effective anti-tumor immunity.

### Contrasting macrophage spatial organization in responders and non-responders

Macrophages are key regulators of the TME and can influence T-cell function. To characterize their spatial organization, CD163⁺ and CD163⁻ macrophages were quantified and their localization relative to tumor and immune cells was assessed. Overall macrophage proportions were similar between ICI response groups, but their spatial organization differed (**Supplementary Fig. S5A**). In non-responders, both protumor (CD163⁺) and antitumor (CD163⁻) macrophages clustered with their own subtype, forming distinct “niches” with minimal mixing (effect size = −0.0955 and −0.1126, respectively; **Fig. 6A**, **Fig. 6B**, top and middle).

**Figure 6.**
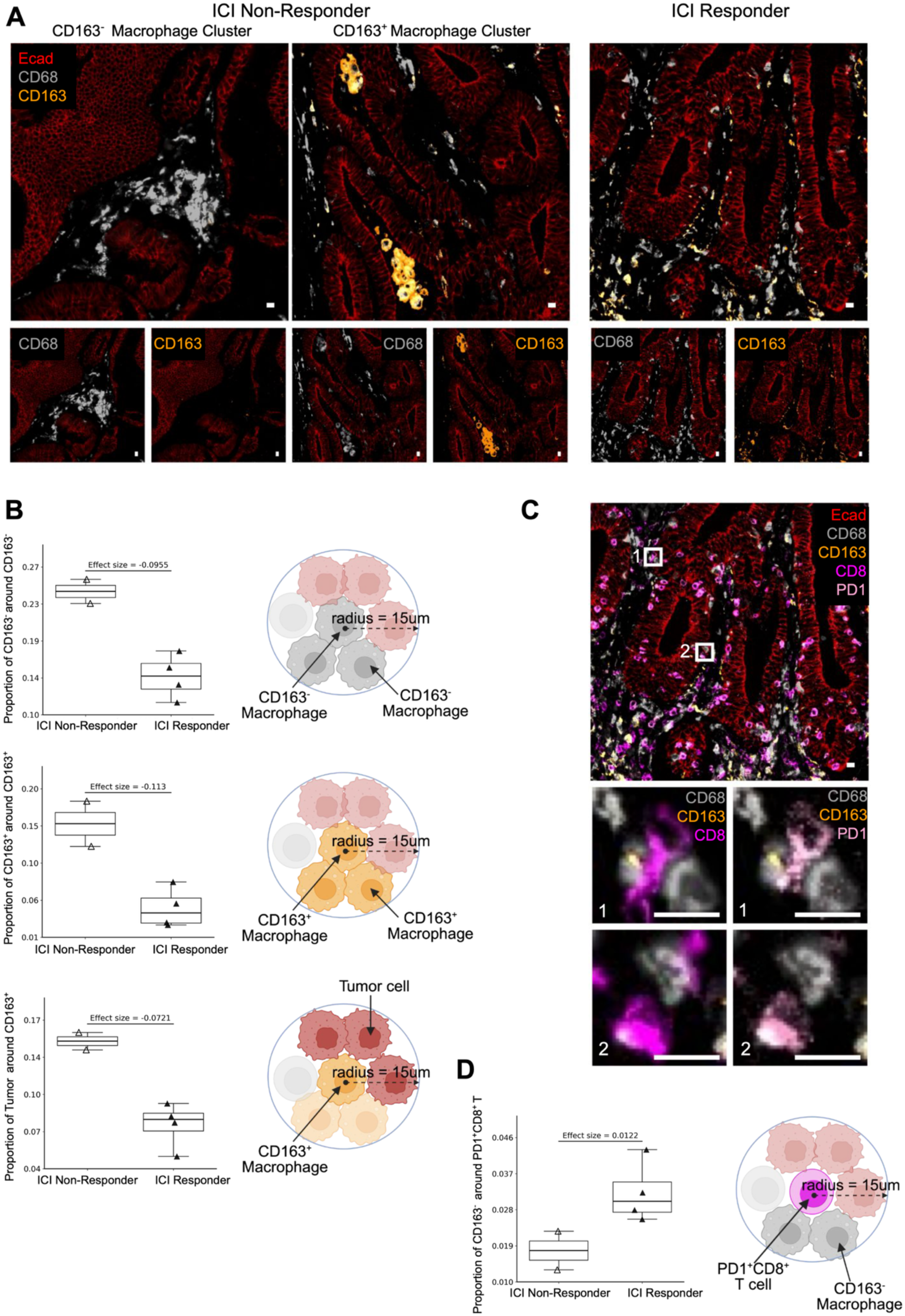
Macrophage spatial organization differs between immune checkpoint inhibition therapy non-responders (n = 2) and responders (n = 4). (A) Representative multiplexed immunofluorescent images illustrating increased self-clustering of CD163^−^ (anti-tumor, grey) and CD163⁺ (pro-tumor, orange) macrophages in non-responder tumors. CD68 and CD163 are shown overlaid (top) and separately (bottom), together with the tumor cell marker E-cadherin (Ecad, red). (B) Mean proportions within a 15 µm radius neighborhood higher in ICI non-responders. Top: CD163^−^ macrophages around CD163^−^ macrophages. Middle: CD163⁺ macrophages around CD163⁺ macrophages. Bottom: Tumor cells around CD163⁺ macrophages. Mean proportion calculated as the number of surrounding cells of interest divided by total cells within the neighborhood and averaged per sample. (C) Representative image showing CD8⁺ T cells (CD8, fuchsia) and CD8⁺ PD-1^+^ T cells (PD-1, light pink) near CD163^-^ macrophages with 2 representative zoomed-in field of views highlighting CD163^-^ macrophages enriched within 15 µm of a PD-1^+^ CD8^+^ T cells (bottom). (D) Mean proportion of CD163^-^ macrophages within 15 µm of PD-1^+^ CD8⁺ T cell calculated as the number of CD163^-^ macrophages divided by total cells in the neighborhood. Due to the small sample size, statistical significance testing was not emphasized; instead, group differences were evaluated based on effect size and the degree of separation (or lack of overlap) between samples.

In non-responders, these CD163⁺ macrophages clustered near tumor cells, with a higher proportion of tumor cells within a 15 µm radius (effect size = −0.0721; **Fig. 6B**, bottom). Similarly, a higher proportion of CD163⁺ macrophages had at least one neighboring tumor cell, and the tumor–CD163^+^ median nearest distance was shorter (**Supplementary Fig. S5B)**.

In responders, macrophage self-clustering was reduced, while interactions between macrophages and T cells increased. CD163^+^ and CD163^-^ macrophages were enriched near CD8^+^ T cells (**Fig. 6C**, top; **Supplementary Fig. S5C**). Most notably, CD163^-^ macrophages were enriched within 15 µm of PD-1^+^ CD8⁺ T cells (effect size = 0.0137, **Fig. 6C**, bottom, **Fig. 6D).** This interaction was consistent across additional spatial metrics, including a higher proportion of PD-1⁺ CD8⁺ T cells with at least one CD163⁻ neighbor and a shorter median nearest distance between PD-1⁺ CD8⁺ T cells and CD163⁻ macrophages in responders (**Supplementary Fig. S5D**). A similar, but less pronounced, enrichment was observed for CD163⁺ macrophages in responders (**Supplementary Fig. S5E**).

These results reveal a divergent spatial TME: in non-responders, both protumor (CD163⁺) and antitumor (CD163⁻) macrophages form dense clusters, with protumor macrophages closely engaging tumor cells, whereas in responders, antitumor macrophages are positioned near PD-1⁺ CD8⁺ T cells, potentially enabling a more successful antitumor response upon ICI therapy.

## DISCUSSION/CONCLUSION

Spatial profiling of dMMR endometrial tumors revealed differences in T-cell composition, macrophage spatial organization, and neighborhood interactions associated with both recurrence and ICI response. Non-recurrent tumors showed higher intratumoral CD8⁺ T-cell proportions and PD-1^Low^ CD4⁺ T cells, whereas recurrent tumors exhibited increased CD4⁺ T-cell proportions and perivascular CD8⁺ T-cell localization. Among ICI-treated patients, responders were enriched for proliferative, antigen-experienced PD-1⁺Ki67⁺ CD8⁺ T cells, and macrophages were more dispersed, with antitumor CD163⁻ subsets frequently co-localizing with CD8⁺ and PD-1⁺ CD8⁺ T cells, forming immune niches associated with therapy responsiveness.

Consistent with prior studies linking infiltrating CD8⁺ T cells to improved outcomes in endometrial cancer, we found a higher proportion of CD8⁺ T cells among all intratumoral T cells to be associated with non-recurrence (24–26). Notably, this association was significant only when CD8⁺ T cells were normalized to the total T-cell population, and not to all cells, consistent with evidence that CD8⁺ to CD3⁺ ratios better reflect functional and clinical relevance (27). Furthermore, this relationship was observed exclusively within the intratumoral compartment, highlighting the region-specific relevance of CD8⁺ T-cell enrichment and suggesting that T-cell composition, rather than overall cellularity, may be a critical determinant of outcome.

While prior spatial profiling studies in endometrial cancer have described immune-inflamed phenotypes associated with improved survival, they did not stratify patients within molecularly defined dMMR cohorts or directly quantify CD8–tumor distances (19). In a breast cancer study, CD8⁺ T cells located within 20 µm of tumor cells were associated with improved survival (17). Our findings demonstrate that similar spatial relationships are present in dMMR endometrial cancer, supporting the biological relevance of tumor enriched niches around CD8⁺ T cells and the inclusion of spatial metrics alongside compositional features in future prognostic models.

In recurrent tumors, CD8⁺ T cells, including PD-1⁺ subsets, were preferentially located near endothelial cells rather than tumor cells, in contrast to non-recurrent tumors where CD8⁺ T cells were enriched within tumor cell neighborhoods, despite similar overall vasculature between groups. This perivascular retention suggests immune exclusion, with T cells are sequestered in vascular niches and have limited access to the tumor parenchyma. Immune-excluded microenvironments have been associated with relapse in endometrial cancer (28), and perivascular localization of CD8⁺ T cells has been reported in multiple solid tumors as a barrier to tumor infiltration (29). Notably, CD8–endothelial spatial relationships have not been directly quantified in endometrial cancer to our knowledge. Our findings support a model in which tumor-associated vasculature restricts parenchymal entry of both CD8^+^ and PD-1⁺ CD8⁺ T cells in recurrent tumors, raising the possibility that vascular architecture contributes to immune exclusion in dMMR endometrial cancer.

In addition to CD8-associated spatial and proportion differences, recurrent tumors exhibited higher proportions of CD4⁺ T cells, whether relative to total T cells or to CD8⁺ T cells, consistent with prior reports of regulatory T-cell enrichment (17, 22). Notably, we found these associations using total CD4⁺ proportions without subset-specific markers, suggesting that broader CD4⁺ enrichment itself may have prognostic relevance. In contrast, PD-1^Low^ CD4⁺ T cells were enriched in non-recurrent tumors, consistent with recent antigen engagement and ongoing activation rather than terminal exhaustion. Across solid tumors, lower PD-1 expression on CD4⁺ T cells correlates with improved outcomes, including reduced recurrence in melanoma and longer progression-free survival in lung cancer (30,31).

Shifting from recurrence to immunotherapy response, we found that higher proportions of PD-1⁺ and PD-1^High^ CD8⁺ T cells were associated with ICI responsiveness. PD-1 marks antigen-experienced T cells, with expression level providing functional insight. This aligns with a prior study in dMMR endometrial and ovarian tumors treated with nivolumab, which reported that the proportion of PD-1⁺ CD8⁺ T cells among total CD8⁺ T cells was significantly higher in responders (32). Although that study used nivolumab rather than pembrolizumab, both are anti–PD-1 inhibitors, supporting the relevance of PD-1⁺ CD8⁺ T cells as a potential biomarker of ICI response. Previous studies have found that high PD-1 expression is often associated with exhaustion or dysfunction (33,34). These cells are prime candidates for reinvigoration by PD-1 blockade, which could help explain why tumors with higher intratumoral proportions of PD-1⁺ CD8⁺ T and specifically PD-1^High^ CD8⁺ T cells respond more strongly to ICI therapy.

CD8⁺ T cells in ICI responders exhibited higher Ki67 expression, indicating increased proliferative activity within the anti-tumor immune compartment. Although this association has not been reported in endometrial cancer, proliferating Ki67⁺ CD8⁺ T cells have been linked to immunotherapy response in melanoma and lung cancer (35–37), while a multicancer analyses suggest that their prognostic significance varies across tumor types (38). Notably, a population of CD8⁺ T cells co-expressing PD-1 and Ki67 has been shown to expand following checkpoint blockade in head and neck cancer, representing antigen-experienced T cells undergoing proliferative reinvigoration after PD-1 pathway inhibition (39). In contrast to these post-treatment observations, our pretreatment analysis shows that ICI-responding tumors contain elevated PD-1⁺Ki67⁺ CD8⁺ T cells. This population is proliferative, antigen-engaged, and poised to respond to checkpoint blockade. Together, these findings suggest that baseline proliferative activity within the PD-1⁺ CD8⁺ T-cell population may identify tumors with a pre-existing immune response predictive of ICI benefit.

In our study, macrophage spatial organization differed markedly between responders and non-responders. Consistent with prior reports linking increased CD163⁺ TAMs to aggressive features and poorer recurrence-free survival in endometrial cancer (26, 27), tumor cells were enriched near CD163⁺ macrophages in ICI non-responders. In non-responders, CD163⁺ and CD163⁻ macrophages clustered within their own subsets, whereas in responders, macrophages were more dispersed and frequently co-localized with CD8⁺ T cells, including PD-1⁺ subsets targeted by ICI therapy. This pattern suggests that reactivated PD-1⁺ CD8⁺ T cells may engage with CD163⁻ macrophages to promote anti-tumor activity. While low CD163⁺ macrophage densities have been linked to improved ICI outcomes in non-small cell lung carcinoma and melanoma (40,41), macrophage-related findings in endometrial cancer have been variable (42–45), and the spatial organization of CD163⁻ macrophages relative to PD-1⁺ CD8⁺ T cells remains unexplored. Together, these findings indicate that co-localization of PD-1⁺ CD8⁺ T cells with antitumor CD163⁻ macrophages represent an ideal immune niche that may facilitate productive immune activation and therapy responsiveness, highlighting an area that warrants further investigation in endometrial cancer.

Several limitations of this study should be acknowledged. Our sample size was constrained by the recent adoption of ICIs for dMMR endometrial cancer, which only became common in clinical practice around 2020, and by the requirement for a minimum of two years of follow-up to assess recurrence outcomes. The small cohort, particularly when comparing ICI responders and non-responders, precluded formal statistical testing; instead, we relied on effect size to identify biologically meaningful differences. All patients were treated at a single academic medical center, which may limit generalizability; future studies incorporating patients from diverse clinical settings are needed to enhance population heterogeneity and representation. Technical limitations included variable staining quality for several markers, necessitating extensive manual quality control and potentially introducing observer bias. Consequently, some markers with potential clinical relevance were excluded from downstream analyses due to insufficient specificity or poor signal. These challenges are consistent with those reported in other retrospective multiplex imaging studies.

Collectively, our results demonstrate that the interplay of T-cell composition, proliferation, activation state, and spatial organization defines recurrence risk and immunotherapy responsiveness in dMMR endometrial cancer. Despite the limited sample size, these findings highlight the potential of spatial profiling to identify clinically actionable features of the TME. Future studies in larger, multi-institutional cohorts are needed to validate these patterns and determine whether they can be translated into robust biomarkers detectable with routine approaches, such as immunohistochemistry or standard H&E staining. By bridging high-dimensional spatial analyses with accessible clinical tools, this framework may advance precision immunotherapy and improve risk stratification in dMMR endometrial cancer.

## Acknowledgements

This work used computing resources at the NYU School of Medicine High Performance Computing (HPC) Facility. We thank Amanda Lund for discussions and feedback; Luis Chiriboga and the Center for Biospecimen Research & Development (CBRD) for the support in obtaining tumor samples; and the Experimental Pathology Research Laboratory (RRID:SCR_017928), which is partly funded by NIH/NCI 5 P30CA16087. This work was also supported by the T32 NYU Grossman School of Medicine Cell Biology Training Grant.

## Authorship Contributions

Study conception & design: M.H., C.H., C.L., L.K., E.A., L.B., D.F.; Performed experiment or data collection: C.H., C.L., S.S.; Computation & statistical analysis: M.H., A.S., W.L., V.M.; Data interpretation & biological analysis: M.H., C.L., E.A., D.F., Writing—Original drafts: M.H.; Writing—Review & editing: M.H., C.H., A.S, W.L., C.L., V.M., S.S., J.T., T.Z., J.W.W., L.K., L.P., E.A., L.B., and D.F., Supervision: E.A, L.B., D.F.; Administration: E.A, L.B., D.F.

## Data Availability

Images will be made available on the public NYU Langone OMERO server at the time of publication.

## Code Availability

Code used for image processing, cell segmentation, cell phenotyping, and spatial analysis is available at: https://github.com/FenyoLab/EC_codeximaging

## Ethics Approval and Consent to Participate

The study was approved by the Institutional Review Board, with a waiver of patient consent.

## SUPPLEMENTAL FIGURES

**Figure S1.**
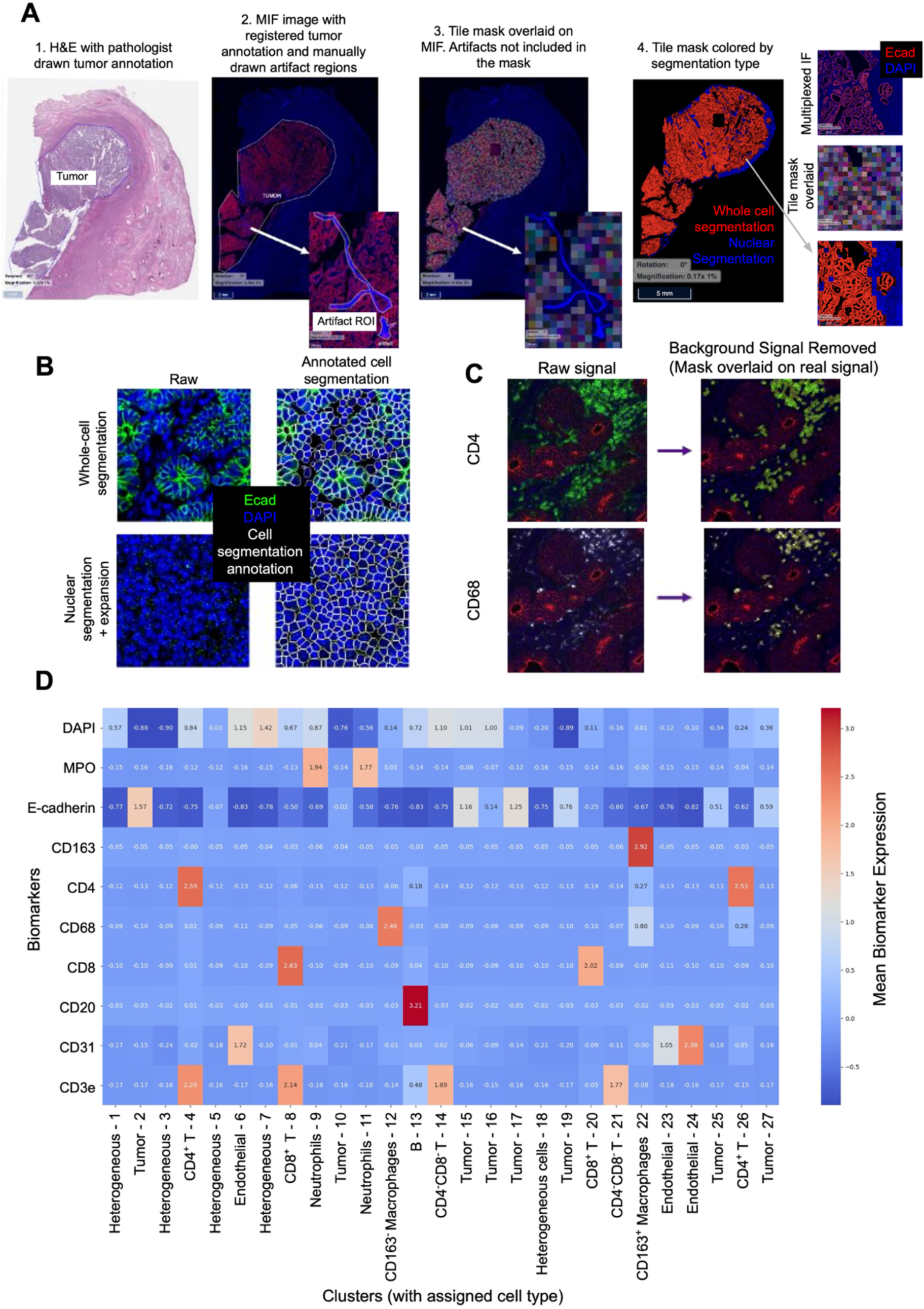
Image preprocessing, cell segmentation, and phenotyping (related to Fig. 2) (A) Pathologist-annotated tumor regions were registered to the multiplex immunofluorescence images. Artifacts were annotated and removed from the tile mask, and each tile was assigned a cell segmentation method. (B) Representative tile showing raw image data (left) and cell segmentation boundaries annotated in white (right) for whole-cell segmentation (top) and nuclear segmentation with 2-pixel (1 µm) expansion (bottom) (green = E-cadherin; blue = DAPI). (C) Marker background removal using k-means clustering (k = 3) illustrated for two example markers (CD4 and CD68), before (left) and after (right) background removal. (D) Clustering results based on lineage markers (k = 27). Each cluster was assigned to a cell type, and heterogeneous clusters were further evaluated to refine cell type assignments.

**Figure S2.**
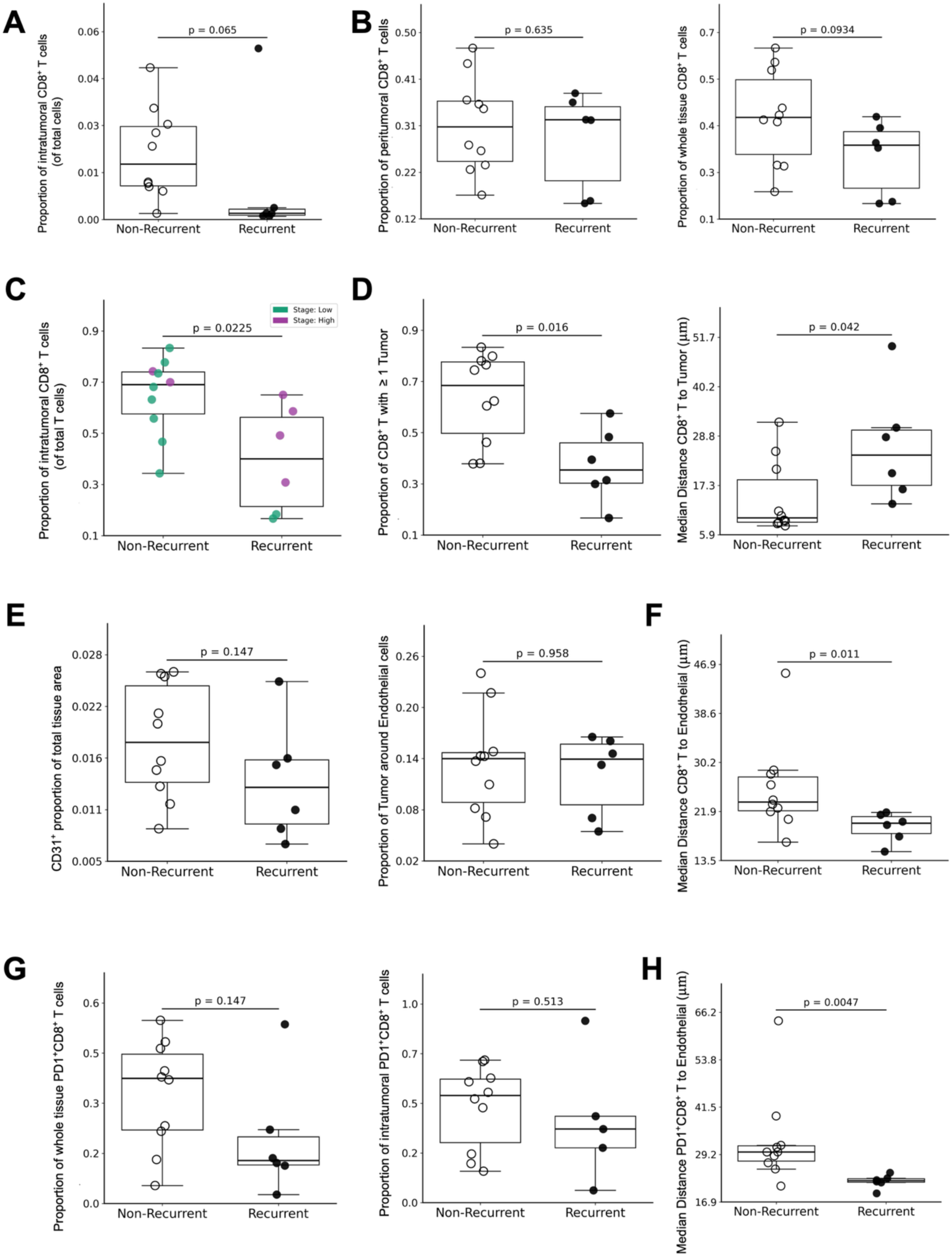
Additional analyses of CD8⁺ T cells and their spatial relationship to tumor and endothelial cells in non-recurrent (n = 10) and recurrent (n = 6) tumors (related to Fig. 3). (A) No significant difference in the proportion of intratumoral CD8⁺ T cells when calculated relative to total cells rather than total T cells. (B) No significant difference in the proportion of peritumoral (left) or whole-tissue (right) CD8⁺ T cells relative to the total T-cell population. (C) Proportion of intratumoral CD8⁺ T cells (of total T cells) stratified by stage (green, low stage [I–II]; purple, high stage [III–IV]). Two low-stage tumors that recurred exhibited the lowest CD8⁺ T-cell proportions. (D) Left: mean proportion of CD8⁺ T cells with at least one tumor cell within a 15 µm radius was higher in non-recurrent tumors. This was calculated as the number of CD8⁺ T cells with ≥1 tumor cell within a 15 µm radius divided by the total number of CD8⁺ T cells. Right: median distance from each CD8⁺ T cell to its nearest tumor cell was shorter in recurrent tumors. (E) No significant differences in vascular metrics between clinical groups. Left: estimated vascular abundance, calculated as CD31⁺ area (mm²) divided by total tissue area (mm²). Right: mean proportion of tumor cells located within 15 µm of each endothelial cell. (F) Median distance from each CD8⁺ T cell to its nearest endothelial cell was shorter in recurrent tumors. (G) No significant difference in the proportion of PD-1⁺ CD8⁺ T cells within the CD8⁺ T-cell population across whole tissue (left), intratumoral regions (right), or peritumoral regions (not shown). (H) Median distance from each PD-1⁺ CD8⁺ T cell to its nearest endothelial cell was shorter in recurrent tumors. *P* < 0.05 by Mann–Whitney U test.

**Figure S3.**
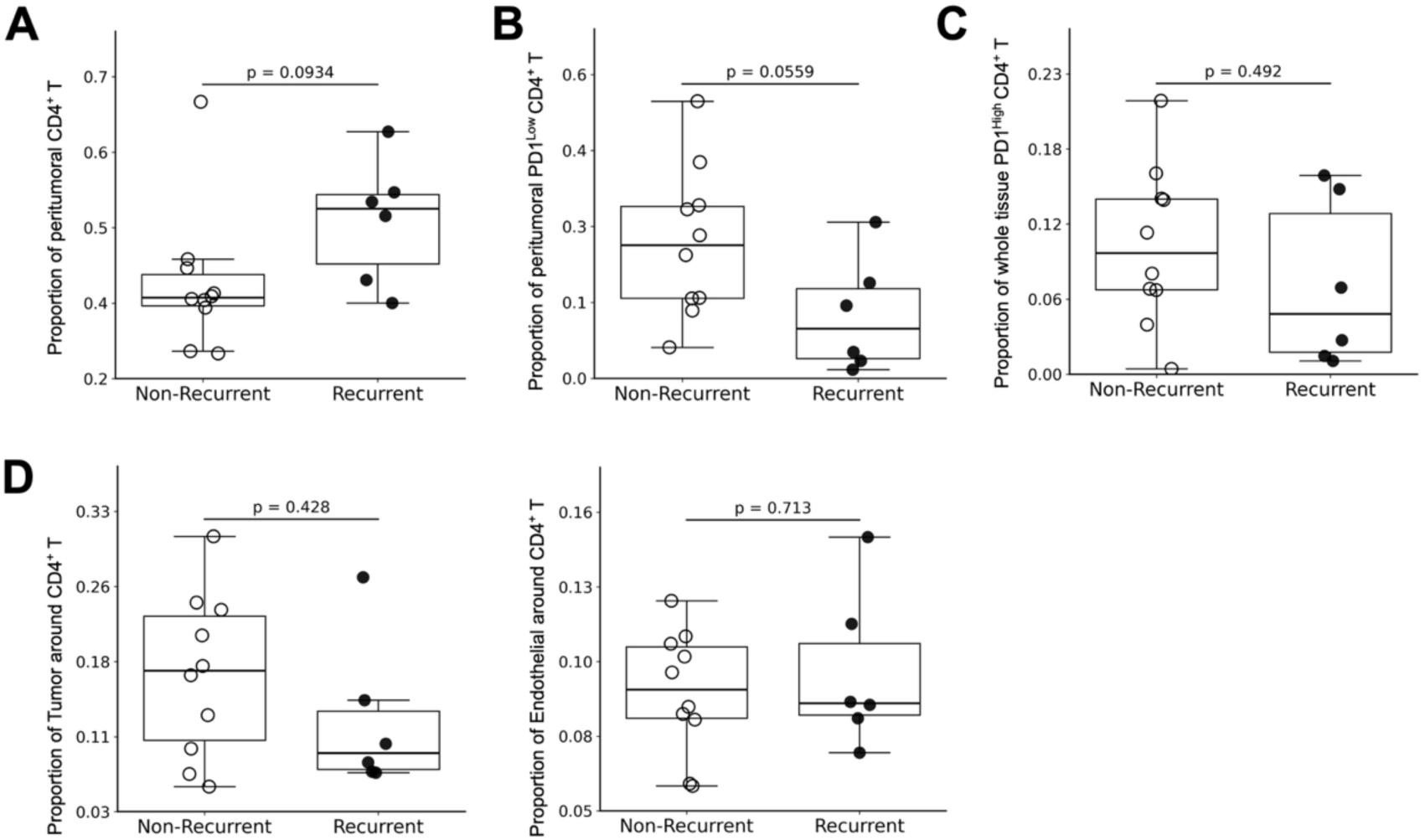
Additional analyses of CD4⁺ T-cell proportions and spatial relationships to tumor and endothelial cells in non-recurrent (n = 10) and recurrent (n = 6) tumors (related to Fig. 4). (A) No significant difference in the proportion of peritumoral CD4⁺ T cells relative to the total peritumoral T-cell population. (B) No significant difference in the proportion of peritumoral PD-1^Low CD4⁺ T cells relative to the total peritumoral CD4⁺ T-cell population. (C) No significant difference in the proportion of PD-1^High CD4⁺ T cells relative to the total CD4⁺ T-cell population across whole tissue. (D) No significant difference in the mean proportion of tumor cells (left) or endothelial cells (right) located within 15 µm of each CD4⁺ T cell. *P* < 0.05 by Mann–Whitney U test.

**Figure S4.**
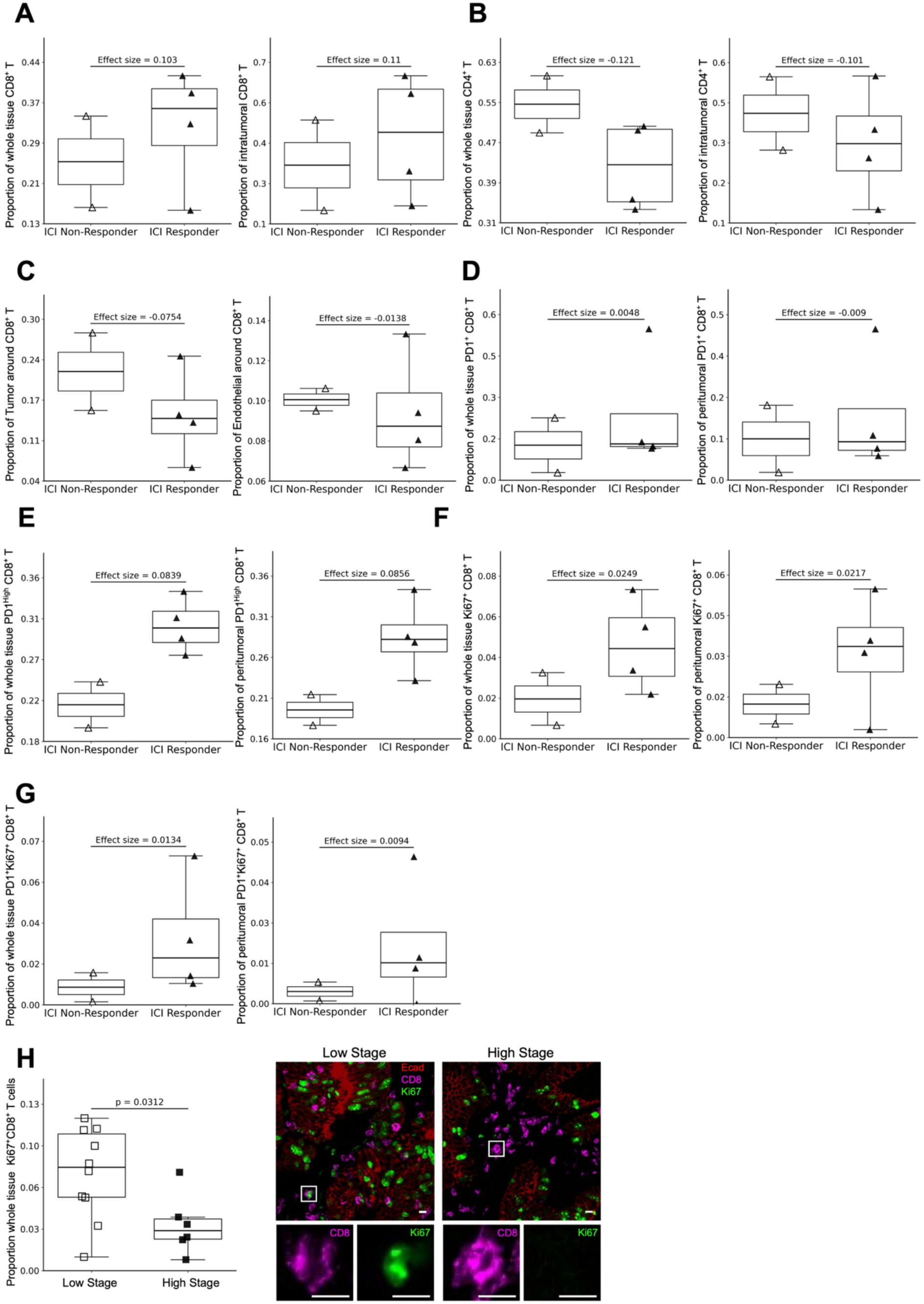
Additional analyses of PD-1⁺ (antigen-experienced) and Ki67⁺ (proliferation marker) CD8⁺ T cells in immune checkpoint inhibition (ICI) non-responders (n = 2) and responders (n = 4) (related to Fig. 5). (A) Proportion of CD8⁺ T cells relative to the total T-cell population in whole tissue (left), intratumoral regions (right), and peritumoral regions (not shown) across response groups. (B) Proportion of CD4⁺ T cells relative to the total T-cell population in whole tissue (left), intratumoral regions (right), and peritumoral regions (not shown) across response groups. (C) Mean proportion of tumor cells (left) or endothelial cells (right) located within 15 µm of each CD8⁺ T cell across response groups. (D) Proportion of PD-1⁺ CD8⁺ T cells relative to the total CD8⁺ T-cell population in whole tissue (left) and peritumoral regions (right) across response groups. (E) Higher proportion of PD-1^High CD8⁺ T cells relative to the PD-1⁺ CD8⁺ T-cell population in ICI responders in whole tissue (left) and peritumoral regions (right). (F) Proportion of Ki67⁺ CD8⁺ T cells relative to the total CD8⁺ T-cell population in whole tissue (left) and peritumoral regions (right) across response groups. (G) Proportion of PD-1⁺Ki67⁺ CD8⁺ T cells relative to the total CD8⁺ T-cell population in whole tissue (left) and peritumoral regions (right) across response groups. (H) Left: higher proportion of Ki67⁺ CD8⁺ T cells relative to the total CD8⁺ T-cell population in low-stage (I–II) compared with high-stage (III–IV) dMMR endometrial tumors (*P* < 0.05, Mann–Whitney U test). Right: representative multiplexed image showing CD8 (fuchsia) and Ki67 (green) co-expression in a low-stage tumor and greater separation of the markers in a high-stage tumor. Scale bar = 10 µm. Effect sizes were calculated as the median in responders minus the median in non-responders. Panels in the main figures highlight features with the largest effect sizes and greatest separation between groups, while additional analyses are presented here for completeness.

**Figure S5.**
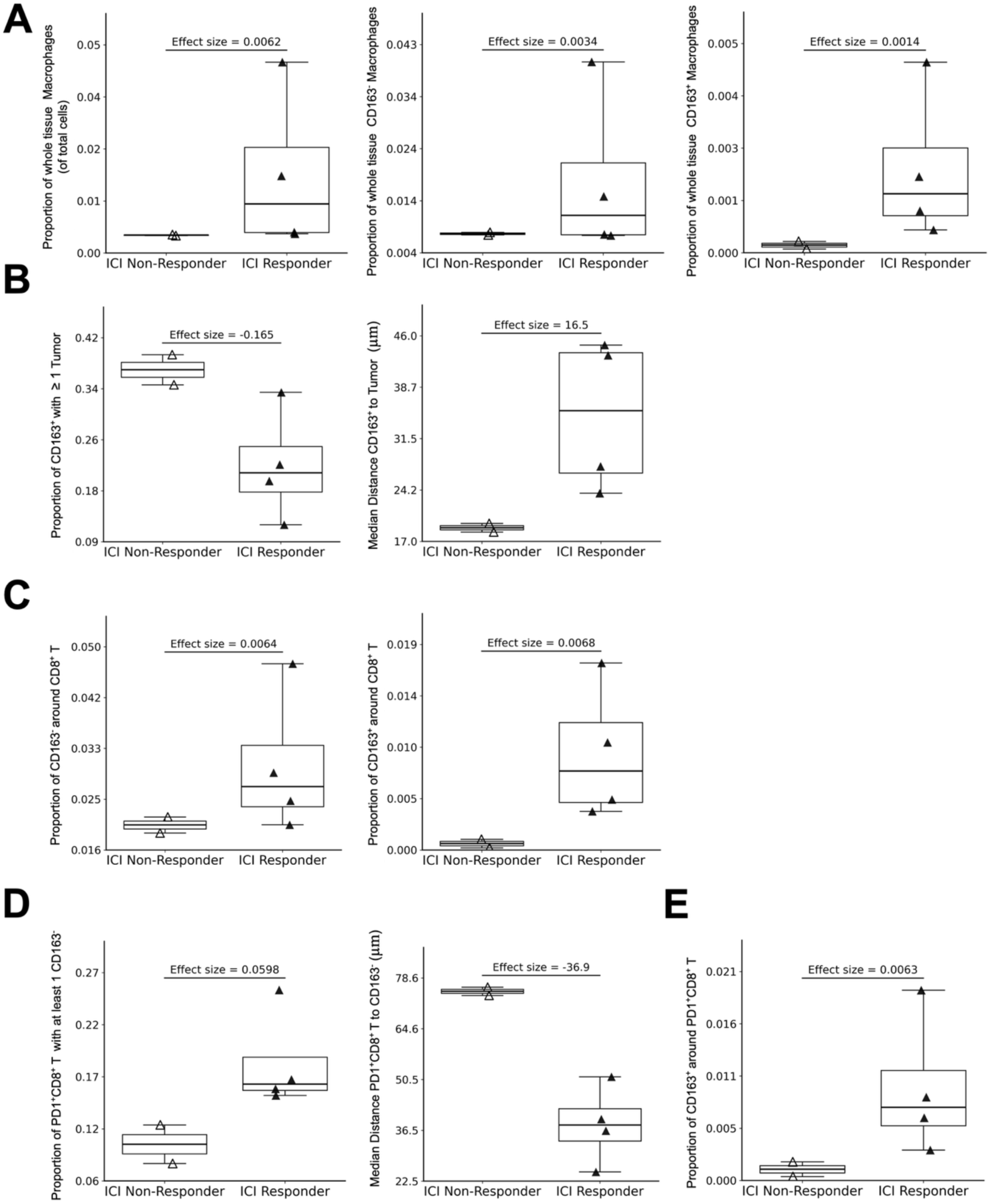
Additional proportion- and spatial-based analyses of macrophages in ICI non-responders (n = 2) and responders (n = 4) (related to Fig. 6). (A) Proportion of whole-tissue macrophages (CD163⁻ and CD163⁺, left), CD163⁻ macrophages (middle), and CD163⁺ macrophages (right) relative to total cells. (B) Left: proportion of CD163⁺ macrophages with at least one tumor cell within a 15 µm radius was higher in non-responders. Right: median distance from each CD163⁺ macrophage to its nearest tumor cell was shorter in non-responders. (C) Mean proportion of CD163⁻ (left) or CD163⁺ (right) macrophages located within 15 µm of CD8⁺ T cells were higher in responders. (D) Left: proportion of PD-1⁺ CD8⁺ T cells with at least one CD163⁻ macrophage within 15 µm was higher in responders. Right: median distance from PD-1⁺ CD8⁺ T cells to the nearest CD163⁻ macrophage was shorter in responders. (E) Mean proportion of CD163⁺ macrophages located within 15 µm of PD-1⁺ CD8⁺ T cells were higher in responders. Effect sizes were calculated as the median in responders minus the median in non-responders. Panels in the main figures highlight features with the largest effect sizes and greatest separation between groups, while additional analyses are presented here for completeness.

**Supplementary Table 1.**
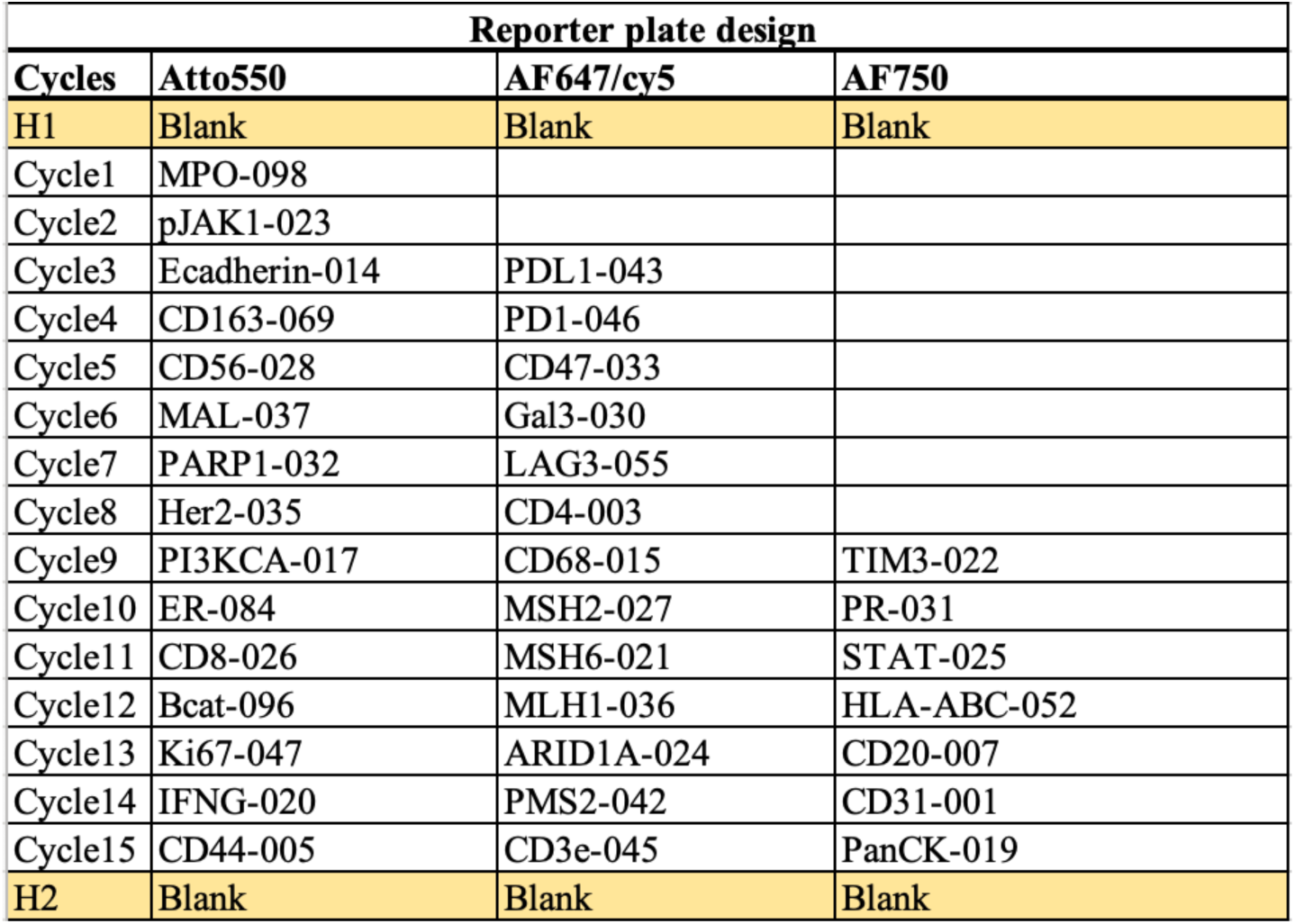
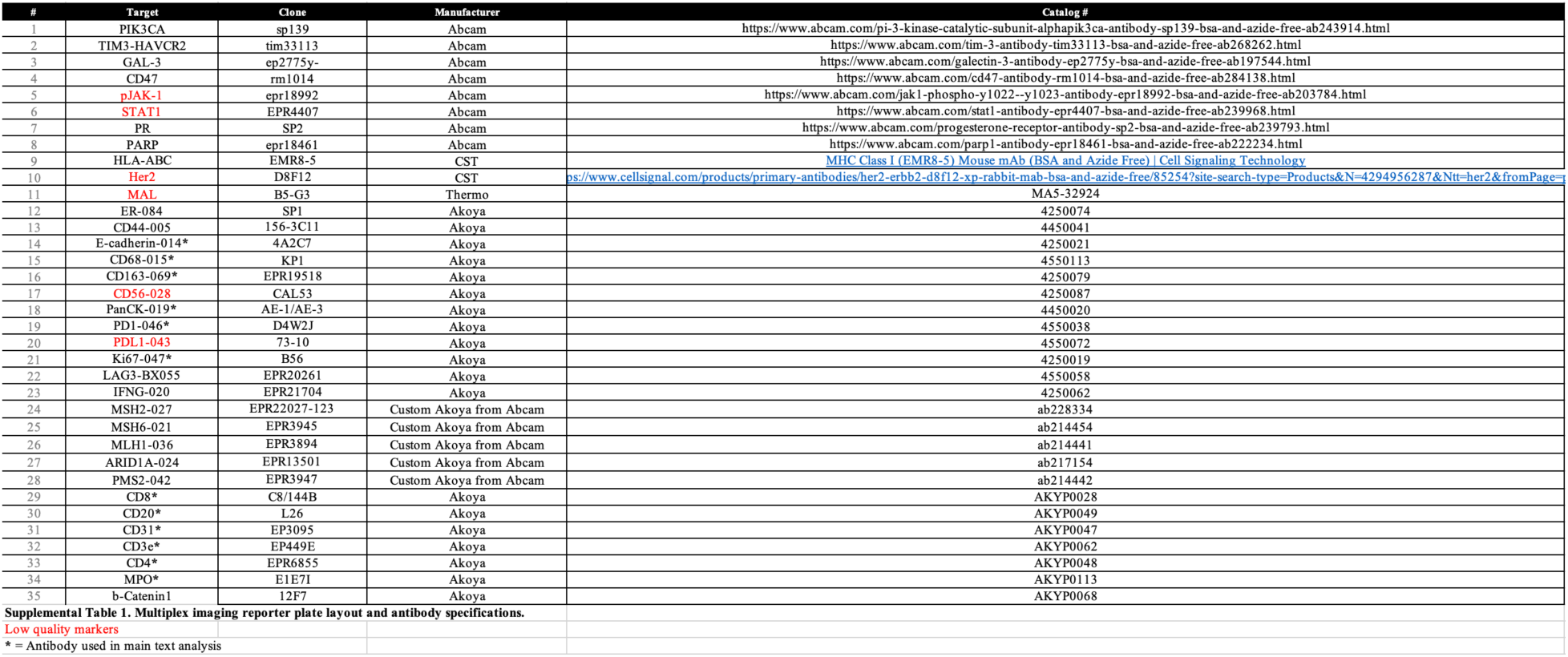
Reporter Plate and Antibody List.

